# Genome-wide SNP genotyping of *Plasmodium falciparum* isolates across Mali reveals major impacts of *Pfsa1* and PfCRT K76T selections on parasite populations

**DOI:** 10.1101/2025.11.11.25339971

**Authors:** Karamoko Niaré, Antoine Dara, Sékou Sissoko, Rebecca Crudale, Aminatou Koné, Hinda Doucouré, Vincent Sanogo, Mady Cissoko, Mahamadou S. Sissoko, Mamadou Tékété, Aissata Koné, Bintou Diarra, Maimouna Dembélé, Boi Koné, Jonathan J. Juliano, Jeffrey A Bailey, Abdoulaye A. Djimdé

**Affiliations:** Department of Pathology and Laboratory Medicine, Brown University, Providence, RI, USA; Center for Computational Molecular Biology, Brown University, Providence, RI, USA; Pathogens genomics Diversity Network Africa, Imm. Gwancoura, Sotuba, Bamako, Mali; Parasites and Microbes Research and Training Center, University of Science, Techniques and Technologies of Bamako, Mali; National Malaria Control Program, Ministry of Health, Bamako, Mali; Institute for Global Health and Infectious Diseases, University of North Carolina, Chapel Hill, NC, USA; Division of Infectious Diseases, School of Medicine, University of North Carolina, Chapel Hill, NC, 27599, USA; Department of Epidemiology, Gillings School of Global Public Health, University of North Carolina, Chapel Hill, NC, 27599, USA

## Abstract

Malaria is a common disease in Mali associated with high morbidity and mortality. To improve malaria control, it is important to evaluate how current interventions affect the genetic make-up of circulating parasites. We analyzed the genomes of recently collected *Plasmodium falciparum* isolates across Mali to understand transmission dynamics, parasite relatedness, and selection pressures affecting parasite populations.

We sequenced clinical isolates (n=458) collected in 2023 from 13 heath districts across Mali using molecular inversion probe (MIP) panels targeting key drug resistance mutations and common SNPs across the *P. falciparum* genomes. We also MIP sequenced polymorphisms in the human β-globin gene in study participants to assess sickle haemoglobin mutation. We performed complexity of infection (COI), identity-by-descent (IBD), principal component (PCA), selection signal, and haplotype analyses.

The prevalence of polygenomic infections ranged from 13.0% in the north, dominated by the Sahara Desert, to 44.0% in the south, with tropical wet and dry climates. Parasites collected from the same district showed low levels of genetic relatedness with a mean IBD of <0.2. There was a network of parasite connectivity between sites across the country, and IBD between districts was inversely correlated with geographical distance separating them (*P=0.006, R=-0.28*). The strongest selection signal was detected around the *chloroquine-resistance transporter* (*pfcrt*) gene. PCA in monogenomic samples (n=242) separated a cluster of parasites (n=56) from the main population showing a specific selection signal around *acetyl-coA synthetase* (*pfacs8)*. The ACS8 Y40F mutation, associated with sickle haemoglobin and also known as *Pfsa1*, was detected in all the samples from this cluster. Higher prevalence of PfCRT K76T mutation (55.8%, n=104) and sickle cell trait (8.4%, n=83) were measured in samples carrying *Pfsa1* compared to wild type (26.6%, n=342 and 1.3%, n=237; respectively, *P<0.005*, Chi-square test). Samples carrying both *Pfsa1* and PfCRT K76T exhibited higher IBD across chromosomes, and one major haplotype was identified around *pfcrt* in 80% of K76T mutants, in which 64% of *Pfsa1* was detected.

Our nationwide genomic profiling of malaria parasites in Mali reveals clear selective sweeps linked to drug pressure and host genetics highly contributing to shaping the genetic make-up of parasite populations, and confirms the association of *Pfsa1* with sickle haemoglobin. The *Pfsa1*–PfCRT 76T linkage highlights a critical axis of parasite adaptation that warrants close surveillance.

## INTRODUCTION

Malaria is a common disease in Mali with significant health and economical impacts. Despite a substantial reduction in malaria burden globally, malaria morbidity and mortality in Mali remain unacceptably high, with ∼8 million cases and ∼20,000 deaths reported in 2022 (*World Malaria Report 2024*, n.d.). Local malaria elimination interventions are currently based on diagnosis, chemoprevention, treatment, and vector control. Artemether-lumefantrine (AL) is the most widely used artemisinin-based combination therapy (ACT) to treat uncomplicated malaria in Mali (*World Malaria Report 2024*, n.d.), although other ACTs, such as artesunate-amodiaquine (ASAQ), are available. While sulfadoxine-pyrimethamine (SP) is used for intermittent preventive treatment of malaria in pregnancy, it is also used in combination with amodiaquine for seasonal malaria chemoprevention (SMC) in children aged between 3 months and 5 years (World Health Organization, 2012). Thus, in Mali, parasites are routinely exposed to five drugs, including sulfadoxine, pyrimethamine, amodiaquine, artemisinins and lumefantrine.

These therapeutic interventions constitute a panoply of selective pressures that can shape the genetic makeup of the circulating malaria parasite populations (Björkman & Bhattarai, 2005; Lubell et al., 2014; Slater et al., 2016). Artemisinin partial resistance has yet to be reported in West Africa despite its emergence and spread in multiple locations along the Rift Valley in Eastern Africa (Conrad et al., 2023; Juliano et al., 2023; Uwimana et al., 2020; Young et al., 2024). Lumefantrine is known to select for the multidrug resistance 1 (MDR1) wild-type N86 allele in populations (Maiga et al., 2021; Vanheer et al., 2023; Veiga et al., 2016). By contrast, amodiaquine selects the MDR1 86Y mutant allele and the chloroquine resistance transporter (PfCRT) K76T mutation (Maiga et al., 2021; Venkatesan et al., 2014). With the widespread use of SP, dihydrofolate reductase (DHFR) N51I, C59R and S108N and dihydropteroate synthase (DHPS) A437G mutations have already approached fixation (Mahamar et al., 2022; MalariaGEN et al., 2023; Vanheer et al., 2023).

In addition to drug pressure, other factors, such as human genetics, unstable transmission, host immunity, and parasite movement, might also affect parasite genetic profiles (Klein, 2013). The most common human genetic polymorphism protecting from malaria is the sickle cell mutation in haemoglobin β-globin gene (HBB) (Allison, 1954). In the heterozygous state (HbAS) or sickle cell trait, it protects from severe malaria by an order of magnitude (Taylor et al., 2012). In the homozygous state (HbSS), it leads to sickle cell disease which is an important public health problem in Mali (Ranque et al., 2022; Sangho et al., 2009). Recently, three variants *–*known as *Pfsa1, 2,* and *3*– in the genome of *P. falciparum* have been positively associated with infections of sickle trait and sickle disease individuals *(Band et al., 2022; Python et al., 2025)*, highlighting important co-evolutionary adaptations in the parasite and human intermediate host.

Genomic analysis has the potential to enhance our understanding of malaria control efforts. By characterizing both parasite and host genetic variation, we can gain insights into parasite population dynamics that would not be revealed by studying the parasite alone. In this study, we investigated transmission dynamics, parasite relatedness and connectivity, population structure, and signatures of selection in parasite populations from 13 health districts across Mali, and examined how these patterns relate to the sickle cell mutation

## RESULTS

### Complexity of infection varies across ecological zones with significant reduction over time in south-western Mali

After MIP sequencing 810 representative samples collected during cross-sectional visits in 13 health care settings across Mali in 2023, we successfully sequenced 458 samples using the IBC2FULL molecular inversion probe (MIP) panel targeting 4,264 common SNPs across the *falciparum* genome (Niaré et al., 2025). For downstream population genomic analyses, we kept samples with <20% genotype missingness rate (n=410, **Table S1**). To evaluate malaria transmission dynamics in different ecological zones, targeted SNPs at minor allele frequency (MAF) ≥1% (2,588 SNPs) were used to estimate complexity of infection (COI) at the admin 1 level, including official regions of Kayes, Koulikoro, Mopti, Segou, Sikasso, and Tombouctou. The prevalence of polygenomic infections –proportion of samples with COI > 1 – was very low in northern Mali with 13.0% in Tombouctou (n=23) and high in southern Mali with ∼44% in Koulikoro (n=92) and Sikasso (n=59) (**Fig. 1A**). The prevalence of polygenomic infections was moderate in south-western Mali (34.1% in Kayes, n=164) and in the wetlands of the central region (32.4% in Segou, n=34; 23.1% in Mopti, n=26). To analyze changes in transmission intensities over time by region, we leveraged the Pf7 whole genome dataset for Malian isolates (n=1,159) collected in 2007 and between 2012–2017 for five locations (Bamako, Kayes, Koulikoro, Segou and Sikasso) and MIP sequenced additional samples collected in 2020 (n=34). We estimated continuous COIs in these older samples and compared them to those of 2023. Significant reductions in COI were observed over time in Kayes and Bamako (**Fig.1B**). Marginal reduction was seen in Segou, but the decrease was not statistically significant. Inrecommended Koulikoro and Sikasso in southern Mali, COI remained stable over time. Overall, the median COI was below 2 in all locations across all timepoints (**Fig.1B**).

**Figure 1:**
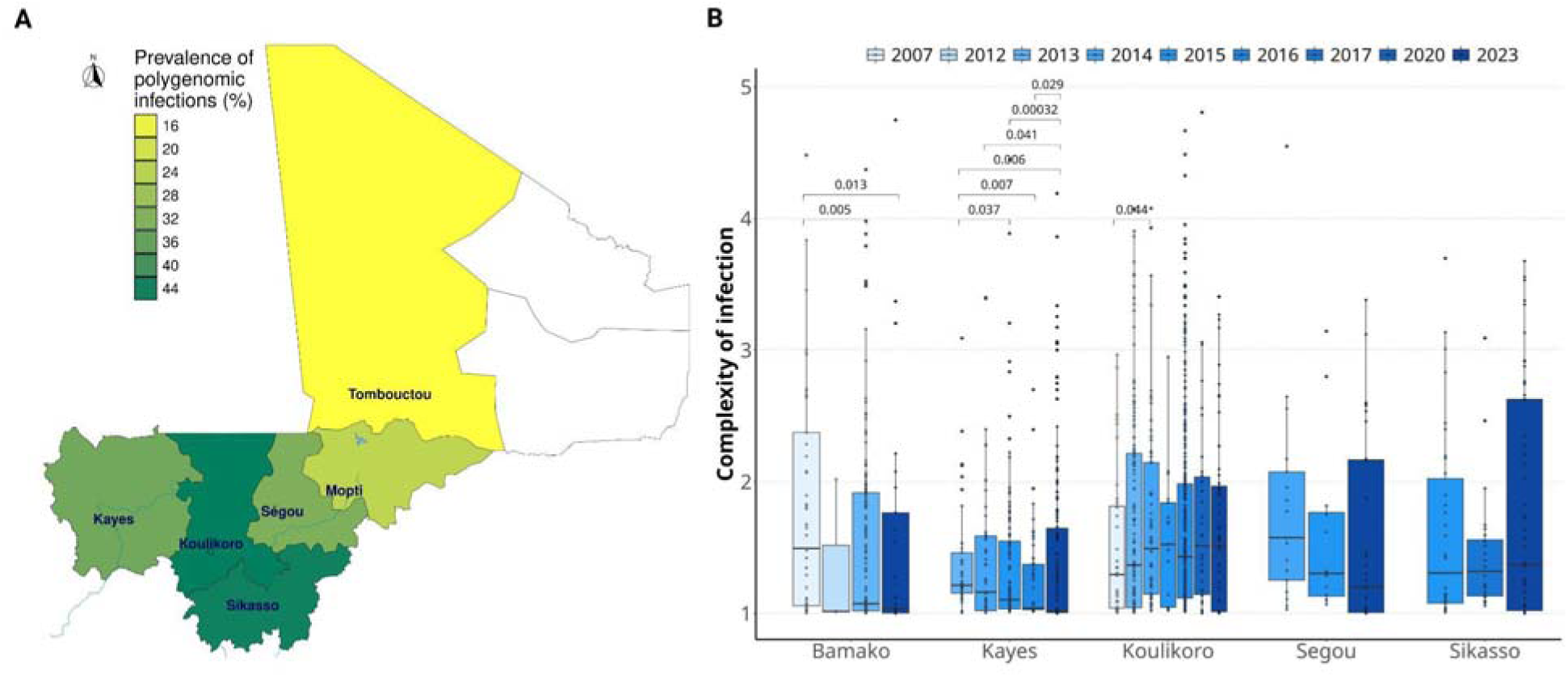
Polygenomic infections across Mali. **A)** Prevalence of polygenomic infections by region (admin1). Data analysis included 410 samples collected in thirteen health districts during the 2023 high malaria transmission. Polygenomic infections indicate samples with COI > 1. **B)** Comparison of complexity of infection over time for five locations. Pf7 dataset was used to calculate COI from 2007 to 2017.

### Pairwise IBD analysis reveals high parasite connectivity across Mali dependent on distance separating sites

To understand recent parasite relatedness across Mali, we calculated pairwise IBD between samples from the same district based on targeted IBC2FULL SNPs at MAF ≥1% and plotted the average values on the map (**Fig. 2A**). We excluded parasite pairs with less than 200 informative SNPs between them. The most closely related parasite populations were observed in southern and northern Mali, with a mean IBD > 0.1. The lowest IBD (0.01 – 0.03) was observed in Mopti along the inner delta of river Niger, a region affected by armed conflicts for a decade. Mean IBD ranged between 0.08 and 0.05 in the rest of the country. To evaluate parasite movement within Mali, we computed pairwise IBD between health districts, and between the precise locations of sample collection. We excluded sample pairs with IBD < 0.25, to track highly related parasites. High IBD sharing was observed between districts that are geographically close to each other(**Fig. 2B**). Three districts (Bamako, Gao and Kidal) studied showed no notable pairwise parasite connectivity (IBD ≥ 0.25) with another location within the country. The strongest connection (Mean IBD = 0.52) was observed between Dire and Taoudeni in northern Mali, suggestive of active local parasite movement in the Sahara desert. There was a significant inverse correlation (*P=0.006, R=-0.28,* Pearson’s correlation) between pairwise IBD and geographic distance between sampling locations, indicating relatedness by distance due to parasite movements (**Fig.2C**). To identify any potential highly related parasite networks present in the country, we analyzed the IBD network at a pairwise IBD ≥ 0.90. Sampling locations were categorized into south, south-west, central, and north (**Table S1**). We detected a large cluster of parasites from multiple locations, primarily from the south and south-west regions (**Fig.2D)**, indicative of active parasite movements between different ecological zones in Mali.

**Figure 2:**
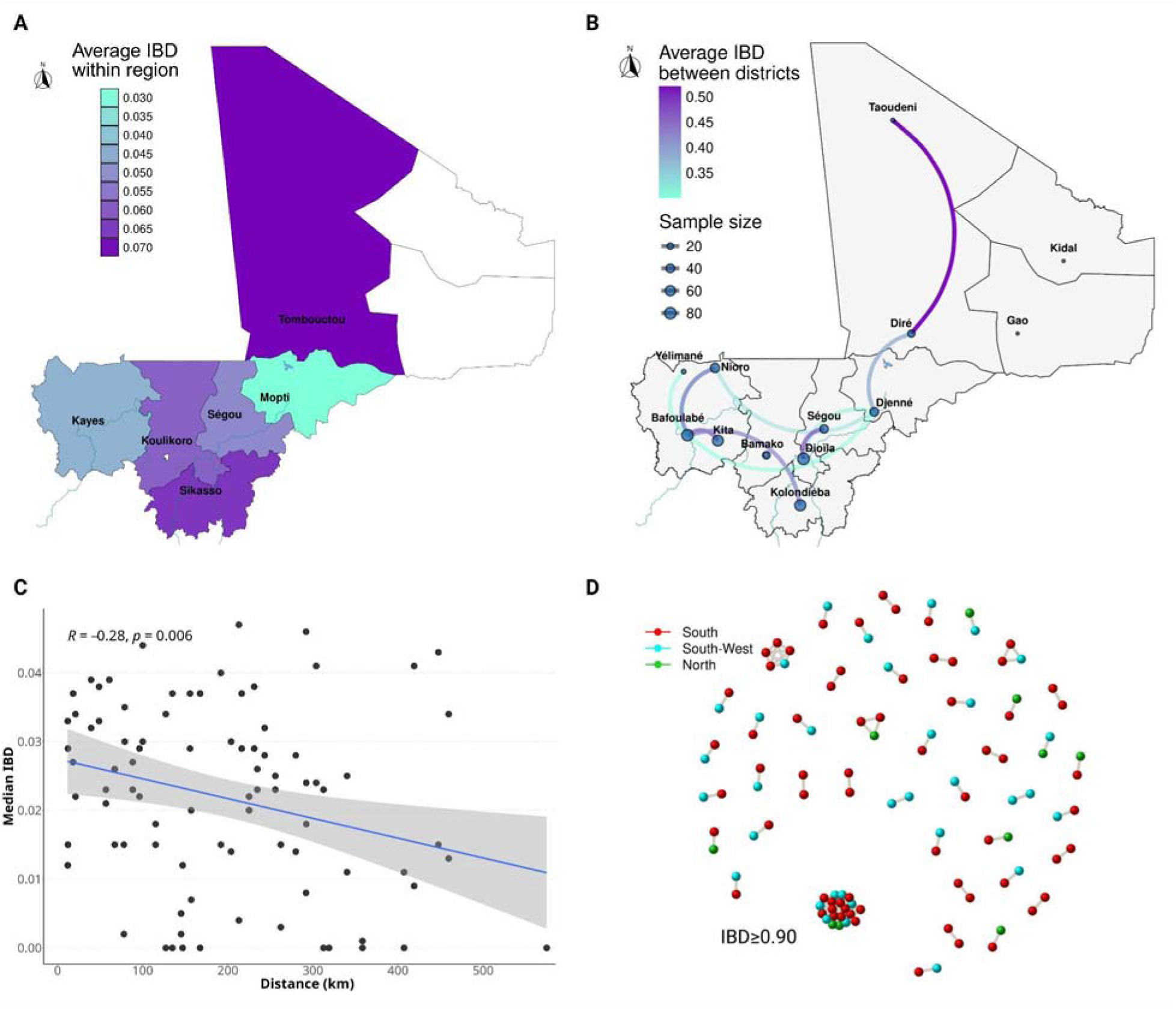
Parasite relatedness and connectivity in 2023. **A)** Pairwise identity-by-descent (IBD) between samples of the same region (n=410). **B)** Parasite connectivity across Mali at IBD at IBD ≥ 0.25. **C)** Inversion correlation between IBD between sampling locations and distance (in km) separating them. Correlation coefficient and *P-*value are indicated. **D)** High IBD network showing a large clonal cluster of parasites. Each dot represents a sample and colors indicate sampling regions.

### Absence of population structure based on SNPs and multiple signals of directional selection

Population structure was analyzed to understand the extent and impact of parasite connectivity across the country. The same SNP set selected for IBD analysis was used to perform principal component analysis (PCA) after correcting for high linkage disequilibrium (r2 <0.2). There was no detectable population differentiation defined by sampling location. Parasites from southern and northern Mali, which had a higher within-district IBD, did not separate from the rest of the population, indicating potential gene flow across Mali (**Fig3.A**).

To detect strong genomic signals of positive selection pressures that might affect these parasites beyond geography, we estimated *IsoRelate* package’s selection metric, iR, across the genome. Using all the samples combined regardless of sampling site (n=410), we detected five selective sweeps after correcting *P-values* for multiple testing by false discovery rate, including *pfcrt, acetyl-coa synthetase* genes *pfacas* and *pfacs8, atp-dependent rna helicase (pfddx60), and DNA-directed DNA polymerase II subunit (pfrpb1)* (**Fig. 3B**). The analysis was also stratified by sampling location, and only districts with ≥ 23 samples were selected, which excluded Gao, Kidal, and Yelimane districts. Samples from Taoudeni (n=4) were merged with those of Dire (n=19) because these parasites showed strong relatedness. We found significant signals (P<10^-5^) in 10 regions of the genome dominated by loci encoding antigens across the 9 districts analyzed (**Fig.S1**). The *pfcrt* locus remained the most common and strongest signal across districts. District-specific signals included *phosphoglucomutase-2 (pfpgm2*), *(pfmsp4)*, *PF3D7_0825900* and *apical merozoite antigen 1 (pfama1*).

**Figure 3:**
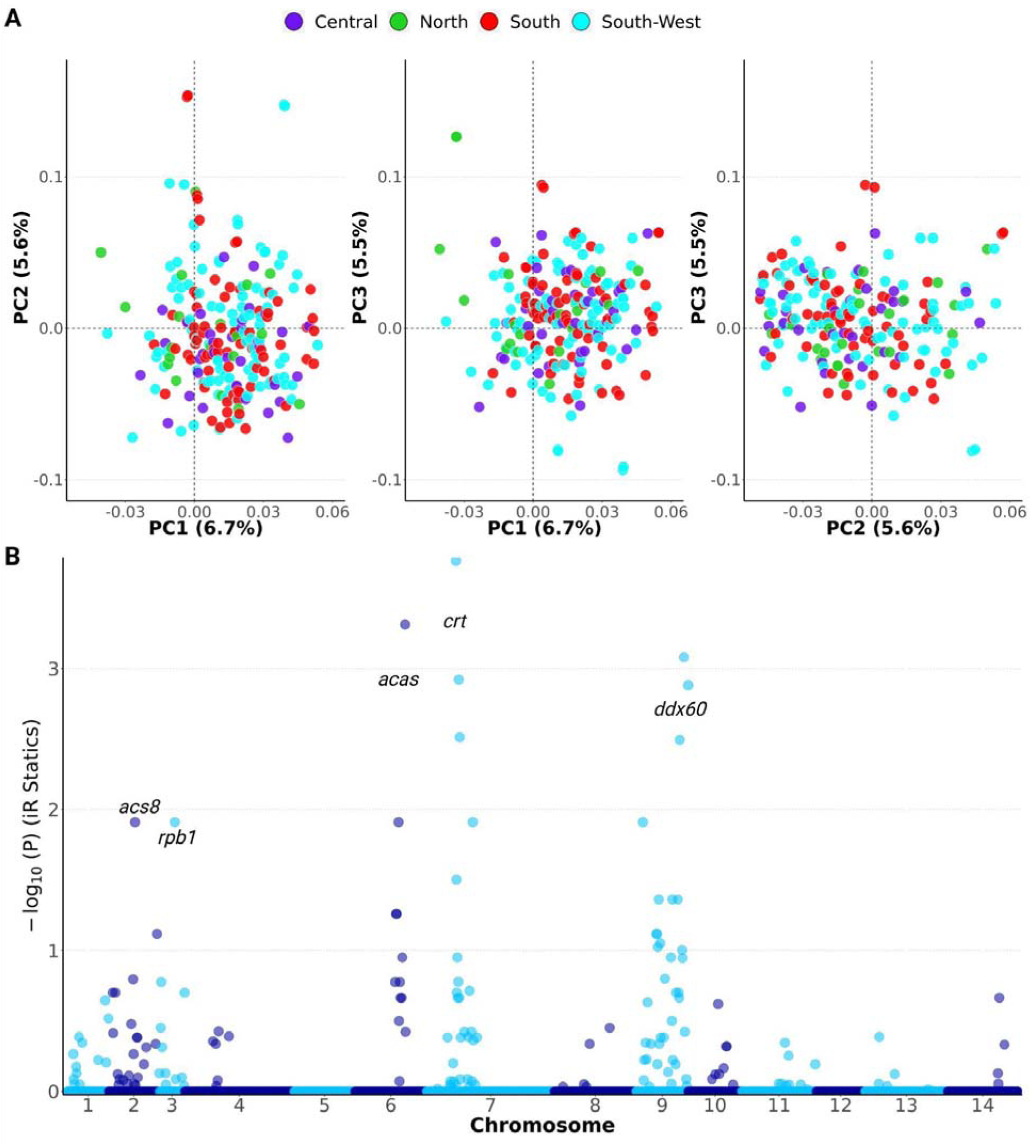
SNPs-based population structure analysis and positive selection signatures. **A)** Principal component analysis (n=410). The first three dimensions (dim1, 2 and 3) are shown and the percentage of variance explained by each of them is indicated in parentheses. Each small dot represents a sample. Large dots represent multiple samples with the same dimensions. Colors indicate sampling regions. **B)** iR selection signals across the genome (n=410). Y-axis represents the *P*-value of iR analysis corrected for multiple testing corrections based on the false discovery rate. Genes found in the iR peaks are indicated.

### Microhaplotypes-based PCA identifies a cluster of *Pfsa1*-carrying parasites associated with PfCRT K76T and sickle trait

Given the population discrimination power of microhaplotypes that has been described elsewhere with closely related parasites (da Silva et al., 2023; Tessema et al., 2022), we reanalyzed population structure based on microhaplotypes. For this analysis, we selected only monogenomic samples (n=242) and extracted microhaplotypes corresponding to the IBC2FULL MIP capture regions (∼100bp) from the variant call format (VCF) file. An F_ST_-based genetic distance matrix based on the presence or absence of the detected microhaplotypes at the MIP capture regions by sample was generated. This was filtered, removing samples and microhaplotypes with 10% and 20% missingness rates, respectively. After performing PCA on the genetic distances, a small cluster (designated group 2) containing 23.6% (57/242) of samples was clearly differentiated by principal component 2 from the main population (designated group 1), independently of sampling locations (**Fig.4A, Fig.S2**). To identify genetic loci contributing to this population differentiation, we scanned the genome for regions that are exclusively identical by descent among parasites belonging to the same population using the iR metric with stratification by population. Five regions were identified in the main population (group 1) corresponding to the regions already detected with the iR analysis based on the entire population or with stratification by district (**Fig.4B**, **Fig.3B, Fig.S1**). Only two signals were identified in group 2 parasites; including *pfcrt*, also present in group 1, and *acs8,* which was absent in group 1 (**Fig.4B**). A previous study has demonstrated that a SNP (Y40F) in ACS8, named *Pfsa1*, is associated with *P. falciparum* evolutionary adaptation to sickle haemoglobin (Band et al., 2022). We evaluated the *Pfsa1* genotypes of our samples, as our IBC2FULL MIP panel already covers this SNP. Interestingly, all the samples from group 2 carried *Pfsa1,* and all the samples from group 1 had the wild-type genotype (**Fig.4C**). To get PfCRT genotypes, the other signal detected in group 2, we sequenced our samples using the existing drug resistance MIP panel, which covers the entire gene (Aydemir et al., 2018; Verity et al., 2020). Additionally, we sequenced polymorphisms in the human HBB gene to analyze the sickle status of the participants upon ethical approval for human genetic studies. The PfCRT K76T mutation was present in both populations (**Fig.S3**) but more abundant (*P=0.015*, Fisher’s Test) in group 2 (47.7%, n=57) than group 1 (30.3%, n=185). We analyzed the prevalence of PfCRT K76T mutations and sickle haemoglobin in samples carrying either PfACAS8 *Pfsa1* or the wild-type genotype using the entire sample set (n=446, including mixed infections). The PfCRT K76T mutation was significantly more prevalent (*P=0.00049,* Chi-square test) in *Pfsa1* genetic backgrounds (55.8%, n=104) compared to ACAS8 wild-type samples (26.6%, n=342) (**Fig.4C**). The same trend was seen when analyzing the PfCRT K76T mutation and *Pfsa1* in samples from Kayes in 2015 (n=154; *P=0.0039,* Chi-square test) and Koulikoro in 2016 (n=427; *P=0.0049,* Chi-square test) using the Pf7 dataset (**Fig.S4**). Examining human, sickle cell trait (HbAS) which was more prevalent (*P=0.0047,* Chi-square test) in participants carrying *Pfsa1* infections with 8.4% (7/83) versus 1.2% (3/237) in those infected by the wild-type samples (**Fig.4C**).

**Figure 4:**
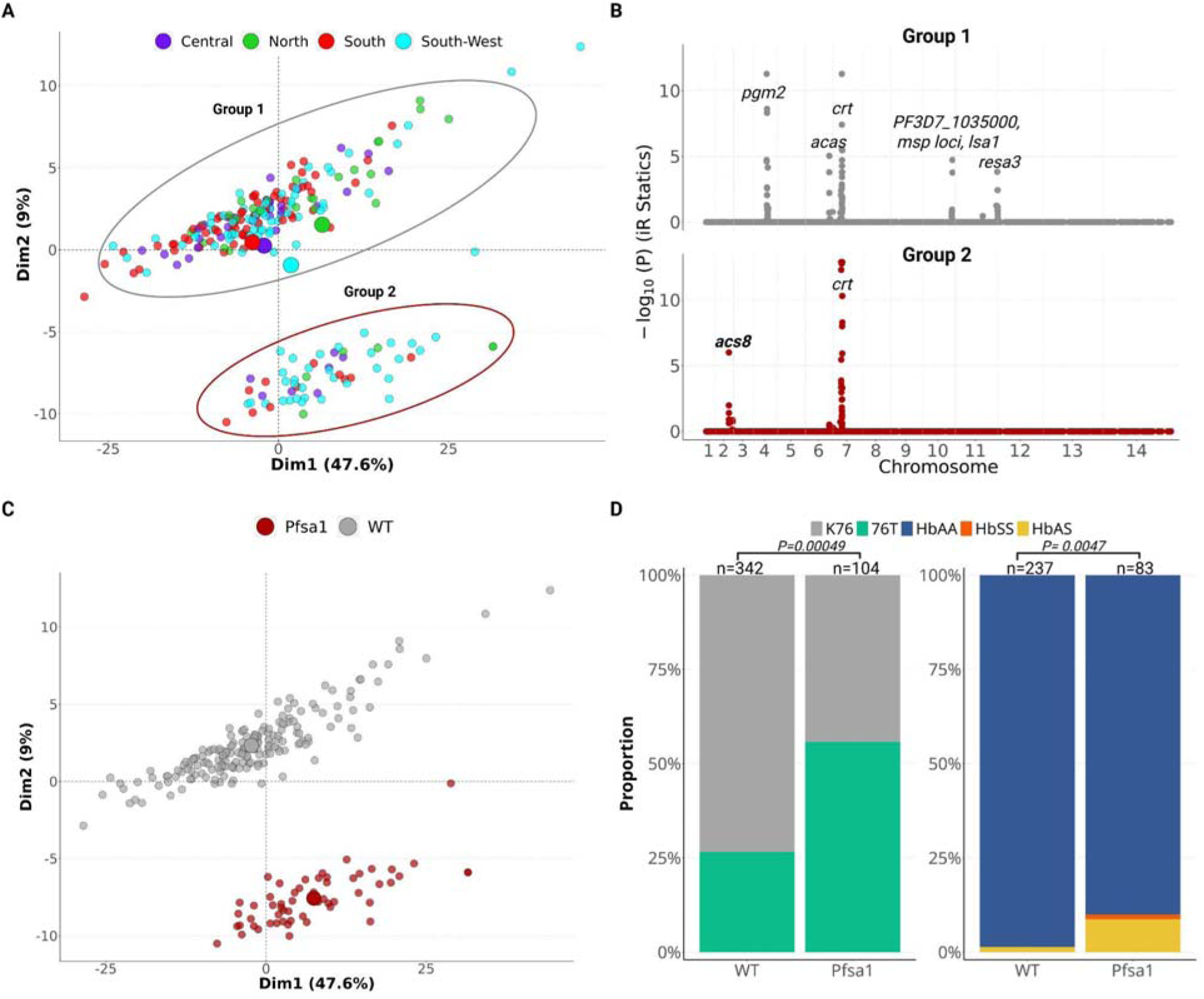
Impact of *Pfsa1* on population structure and its association with the PfCRT K76T mutation and sickle haemoglobin. **A)** Microhaplotypes-based PCA in mono-genomic samples (n=242) colored by sampling regions. First two dimensions (dim1 and 2) are shown and the percentage of variance explained by each of them is indicated in parentheses. The two groups of parasites (1 and 2) differentiated are highlighted by gray and red ellipses, respectively. **B)**. The iR scan for selection signals in group 1 (n=185) and 2 (n=57). Each dot represents a SNP. Y-axis represents *P-*value corrected for multiple testing based on false discovery rate. Genes found in the iR peaks are indicated. **C**) PCA identical to A) but with samples colored by their *Pfsa1* genotype. WT: wildtype. **D)** Comparison of the prevalences of PfCRT K76T mutation and sickle cell trait in samples carrying *Pfsa1* versus wildtype. Sample sizes and *P-*values (Chi-square test with Monte Carlo simulation using 100,000 replicates) are indicated. HbAA: normal haemoglobin in both chromosomes. HbSS: homozygous for sickle haemoglobin, HbAS: heterozygous carrier of sickle haemoglobin or sickle cell trait.

Although a marked population structure was not found with PCA based on SNPs (**Fig3A**), *pfacs8* and *pfcrt* loci were among genomic regions showing high loading values (**Fig. S5**).

### Higher IBD and one major *PfCRT* haplotype detected among parasites carrying both *Pfsa1* and PfCRT K76T mutation across Mali

To understand the impact of the selection pressures directed toward *Pfsa1* and PfCRT K76T mutation on parasite populations, we compared IBD from sample pairs carrying both genotypes with that from sample pairs with either only one of the alleles or the wild types. These IBD comparisons were performed across the fourteen chromosomes using the IBC2FULL data. As expected, the highest IBD sharing was observed for chromosomes 2 and 7, which carry the *pfacs8* and *pfcrt* genes, respectively. However, samples carrying *Pfsa1* and the PfCRT K76T mutation also had higher IBD across the other chromosomes compared to other genotypes. Significant sharing was found for chromosomes 2, 5, 6, and 7 (**Fig. 5A)**; suggesting a shared genetic background among isolates with both genotypes. To examine the evolutionary history of these genotypes in Mali, we analyzed haplotype patterns around the *pfacs8* and *pfcrt* genes in 130 monogenomic samples randomly selected from different sites. Although our sequencing did not tile the entire flanking regions of these genes, our MIP panel captured 118 and 16 SNPs within 45Kb and ∼300kb regions around *pfcrt* and *acs8*, respectively, for haplotype pattern analysis. For the *pfacs8* gene, all *Pfsa1* samples with or without the PfCRT K76T mutation clustered together and showed 4 noticeable haplotypes with signs of extensive recombinations in the flanking regions (**Fig.5B**). For *pfcrt*, we detected one major long haplotype representing 80% (64/80) of K76T samples in which most of (64%, 22/34) *Pfsa1* mutants belong (**Fig.5B**), indicating that the vast majority of K76T mutations currently spreading in Mali derived from one common line whose genetic background likely contained *Pfsa1.* Eleven other minor K76T haplotypes, including 20% (16/80) of K176T samples were detected, each containing 1-6 samples, in which only 11% (4/34) of *Pfsa1* were found (**Fig.5B**).

**Figure 5:**
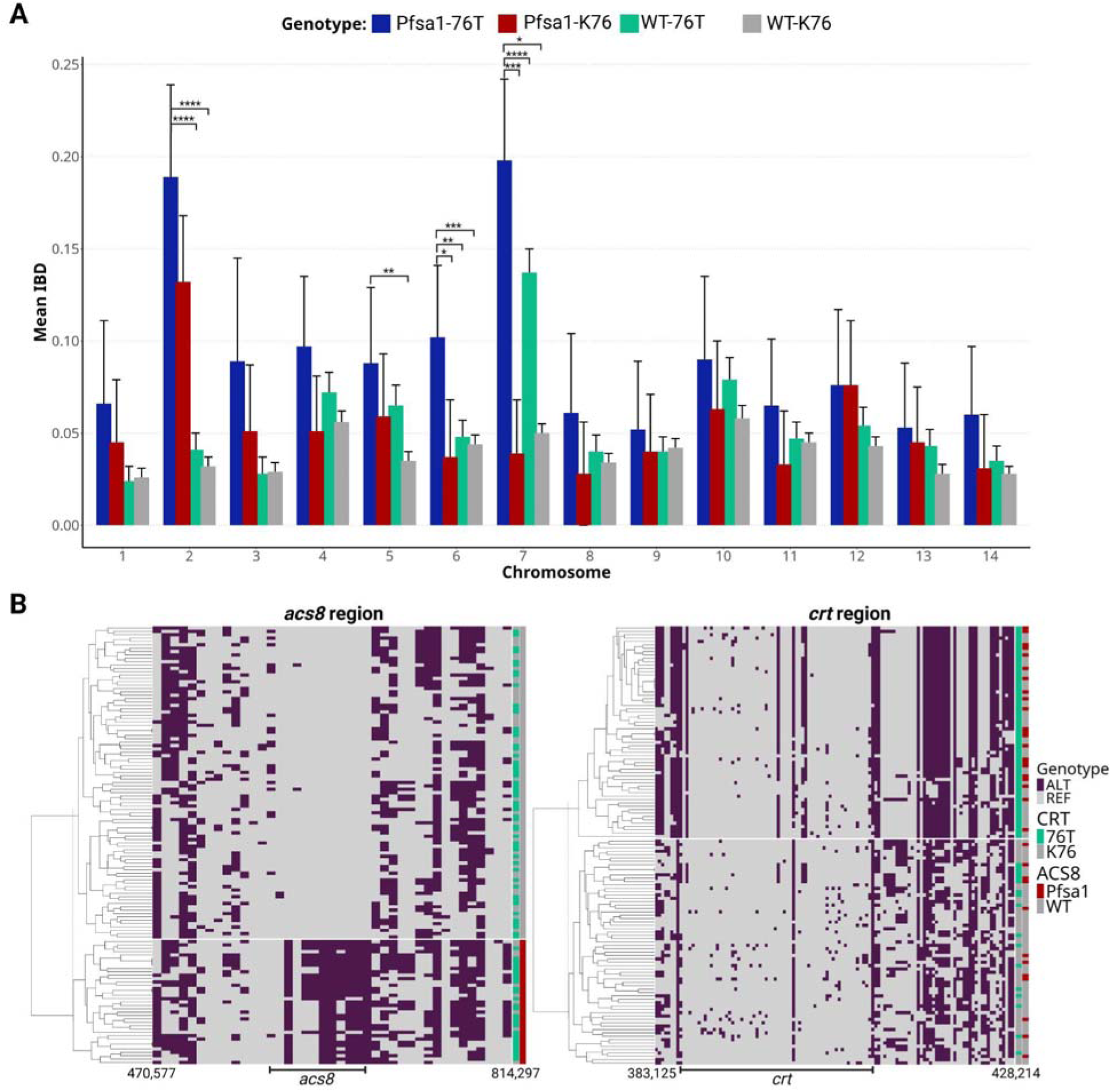
IBD profile and haplotype patterns of samples carrying *Pfsa1* and the PfCRT K76T mutation. **A)** Chromosome-level IBD from sample pairs carrying both *Pfsa1* and the PfCRT 76T mutant (*Pfsa1*-76T) versus those from sample pairs with *Pfsa1* and the K76 allele (*Pfsa1*-K76), wild-type ACS8 and 76T (WT-K76) and wild-type ACS8 and K76 (WT-K76). Levels of significance are indicated by asterisks. 410 samples were used. **B)** Visualization and clustering of haplotypes flanking *acs8* and *pfcrt* in monogenomic samples (n=130). 42 SNPs at MAF ≥1% within a ∼300kb region were analyzed for *pfacs8.* 119 SNPs at MAF ≥1% within a ∼45kb region were analyzed for *pfcrt*.

## DISCUSSION

Analyses of COI, parasite relatedness, population structure, and selection signals are complementary to drug resistance assessment in implementing molecular surveillance of malaria in endemic regions. This study provided detailed information on the genomic epidemiology of malaria across all ecological zones in Mali, a large country in West Africa with 1,240,192 square kilometres, after sequencing common SNP barcodes across the *P. falciparum* genome using the MIP technology. Our findings are representative of recent malaria parasite populations in Mali, as the study includes samples collected in 2023 from thirteen different districts. We found differences in COI and parasite relatedness between regions. We demonstrated long distance pairwise parasite connectivity across the country as well as a trend supporting genetic isolation by distance. We detected several genomic signatures of positive selection, including two notable regions surrounding the *pfcrt* and *pfacs8* genes that influenced parasite population structure and shared a common genetic background. High prevalences of the PfCRT K76T and PfACS8 Pfsa1 mutations were observed, which have been previously associated with chloroquine resistance (Djimdé et al., 2001) and the sickle hemoglobin trait (Band et al., 2022), respectively. The vast majority of the PfCRT K76T mutations spreading in Mali derived from one major haplotype.

The current selection of the PfCRT K76T mutation seemed to be driven by ongoing interventions based on the use of amodiaquine. The first emergence and spread of this mutation in Mali was historically due to the widespread use of chloroquine (Djimde et al., 2010), but this drug was replaced by ACTs in 2006. Additionally, studies have indicated a high fitness cost of the PfCRT K76T mutation, which gradually disappeared from the population after abandoning chloroquine (Laufer et al., 2006; Liu et al., 1995). Amodiaquine is an amino-4-quinoleine related to chloroquine, currently combined with SP for use in SMC and artesunate for the treatment of uncomplicated malaria (World Health Organization, 2012). The selection of the PfCRT K76T mutation by amodiaquine has been demonstrated with therapeutic efficacy studies, and our findings are in keeping with this evidence (Venkatesan et al., 2014). Whole genome sequencing studies of malaria parasites elsewhere, where amodiaquine is still officially used, also reported strong positive selection signals around the *pfcrt* gene (Amambua-Ngwa et al., 2018; Duffy et al., 2015; Schaffner et al., 2023). Mali has implemented SMC, which consists of systematically giving SP with amodiaquine to children monthly during malaria transmission seasons, in most health districts across the country from 2012 to 2016 (Cissoko, Sagara, et al., 2022; Konaté et al., 2020). In addition to the SMC, more amodiaquine selection pressure is likely generated by the use of ASAQ, the second most widely used ACT in Mali after AL (Diarra et al., 2021).

Another major detected signal of selection pressure affecting population structure was PfACS8 *Pfsa1,* also frequently seen in the genetic backgrounds of parasites containing the major K76T haplotype. The impact of *acs8* on population structure has also been previously described in Tanzania and the Democratic Republic of Congo (Moser et al., 2020; Verity et al., 2020). We detected a higher prevalence of HbAS carriage in individuals infected by parasites with the *Pfsa1* genotype, which is consistent with previous studies demonstrating the association between this polymorphism and sickle hemoglobin (Band et al., 2022; Hamilton et al., 2023). These findings are also consistent with the high prevalence of sickle haemoglobin in Mali (Ranque et al., 2022; Sangho et al., 2009). However, beyond the impact of sickle haemoglobin, we noted a genetic association between *Pfsa1* and PfCRT K76T evidenced by higher IBD sharing among sample pairs carrying both genotypes, which was seen across the entire genome and not only on chromosomes 2 and 7, where these genes are located, respectively. Our haplotype analysis provided further understanding of this association. First, the extensive recombination noticeable around *Pfsa1* is indicative of the ancient nature of the selection pressure directed toward this genotype, which would have predated the emergence or the reselection of the PfCRT K76T mutation. Second, the long shared haplotype flanking the K76T mutation indicated a recent selection or a reselection of this genotype by amodiaquine. Third, the most common and longest PfCRT haplotype, which was found in 80% of K76T-carrying samples in Mali, was also enriched in *Pfsa1.* This suggests that the K76T haplotype likely emerged from a genetic background that already contained *Pfsa1.* The preferential spread of this haplotype compared to others requires further investigation. Conducting the same study elsewhere, where amodiaquine is used and sickle haemoglobin is common, may provide more clarification on the potential role of *Pfsa1* in the spread of the PfCRT K76T mutation.

In addition to PfCRT and PfACS8, other strong selection signals were detected, including PfACAS, PfDDx60, and PfPGM2. Biochemical studies have identified PfACAS as the molecular target of active metabolites derived from pantothenamide (MMV693183), a new antimalarial drug that has undergone preclinical assessments (de Vries et al., 2022). Like PfACS8, PfACAS is also a member of the acetyl-coenzyme A synthetases whose selection can be investigated using whole genome sequencing. PfDDx60 is an ATP-dependent RNA helicase involved in the regulation of stress survival (Tuteja, 2016), and PfPGM2 is a putative phosphoglycerate mutase (Hills et al., 2011). Studies investigating the role of K13 Interacting Candidate 5 protein (PfKIC5) in regulating artemisinin stress response have found downregulation of PfDDX60 in KIC5-disrupted parasites (Simmons et al., 2023).

This study provides new insights into the transmission landscape of malaria across Mali’s diverse ecological zones. The observed variation in the prevalence of polygenomic infections was consistent with previous descriptions of malaria ecology in the country (Cissoko, Magassa, et al., 2022). However, our work represents the first nationwide molecular assessment of the COI, a recognized correlate of malaria transmission intensity (Schaffner et al., 2023). Polygenomic infections were more frequent in southern Mali, characterized by a tropical climate and abundant rainfall, where malaria incidence is high, compared to the arid northern regions dominated by the Sahara Desert, where transmission is sporadic (Cissoko, Magassa, et al., 2022). The intermediate prevalence of polygenomic infections observed in south-western Mali with a semi-arid climate, and in the central wetlands aligns with the moderate malaria incidence reported in these regions (Cissoko, Magassa, et al., 2022). In addition to these profiles, this study demonstrated a significant decrease in the COI in south-western Mali and in Bamako, the largest city of the country, over time; highlighting the local impacts of control interventions on malaria transmission intensity.

Overall, the IBD profile at the regional level reflects the recent history of human population movements across the country. Since 2012, Mali has been affected by armed conflicts in northern and central regions, resulting in massive local population displacements and military activities internally. This intensive population movement is a major factor contributing to the gene flow, generating heterogeneity in parasite populations. Consequently, the lowest within-district IBD was observed in the region of Mopti in central Mali, one of the most affected locations of the country. The human movement further reflects the wide network of parasite connectivity between neighboring districts, the inversion correlation between pairwise IBD between sites and the distance separating them, and the large clonal cluster of parasites from different regions.

As limitations, the analysis included a few samples from health districts in northern Mali due to ongoing armed conflicts, the lower incidence, and that malaria typically occurs as outbreaks in this region. Only ∼50% of the samples sequenced generated data for downstream analyses, due to sample collection based on a rapid diagnostic test and not microscopy. Further investigations of the multiple selections detected were limited by the lack of whole-genome sequencing.

In conclusion, this study provides a detailed insight into the genomic epidemiology of malaria country-wide in Mali and identifies selection signals mainly due to the impact of ongoing use of amodiaquine for chemoprevention and treatment, and sickle haemoglobin highly contributing to shaping the genetic make-up of parasite populations.

## MATERIALS AND METHODS

### Sample collection and study settings

We collected dried blood spot samples (DBS) from individuals tested positive for malaria by rapid diagnosis tests during the 2023 transmission season as part of a country-wide surveillance of hrp2/3 deletions. We used 810 DBS collected during the survey for genomic studies. Additionally, 94 archived samples, collected in 2020 from the health district of Kati in the region of Koulikoro, were used. In total, 13 health districts were included, including Bafoulabe, Kita, Nioro and Yelimane in south-western Mali; Dioila, Kati, Kolondieba in southern Mali; Djenne and Segou in the central Mali; Dire, Gao, Taoudeni and Kidal in northern Mali. These sites represent all ecological zones and the current malaria transmission strata in Mali (Cissoko, Magassa, et al., 2022). South-western Mali has a hot semi-arid climate according to the Köppen climate classification (Beck et al., 2018) with a hot dry season from November to May and a wet rainy season from June to October corresponding to the peak of malaria transmission. Southern Mali is the wettest area of the country where malaria transmission is known to be highest with abundant rainfall due to a tropical wet and dry climate (Cissoko, Magassa, et al., 2022). The central area is dominated by the inner Delta of Niger River, a wetland region with hot semi-arid climate where the peak of malaria transmission occurs during the rainy season from June to October. Northern Mali corresponds to the Sahara Desert, the hottest area of the country with a hot desert climate, rare rainfalls and sporadic malaria epidemics. Ethical approval was obtained from the ethics committee of the Faculty of Medicine and Odontostomatology and the Faculty of Pharmacy of Bamako.

### Sequencing and variant calling

In total, DBS (n=810) were subject to DNA extraction using a Chelex-Tween protocol before MIP capture (Aydemir et al., 2018; Verity et al., 2020). We used different MIP panels to capture common SNPs across the genome (IBC2FULL panel) (Niaré et al., 2025), key drug resistance mutations and polymorphisms in human HBB gene into circular DNA, as previously described (Aydemir et al., 2018; Deutsch-Feldman et al., 2019). After digesting linear DNA with exonucleases, sample barcodes were added to the captured DNA by PCR amplification to create an Illumina library for sequencing on an Illumina Nextseq 550 platform at Brown University. We used laboratory strains 3D7 and Dd2 used as controls in all the experiments. The MIP design contained unique molecular identifiers (UMIs) which were utilized to remove PCR and sequencing errors. We used the MIPTools software suite (version4.0) (https://github.com/bailey-lab/MIPTools) for the data analysis which consisted of extracting and mapping microhaplotypes onto the reference genome and calling variants by Freebayes (version1.3.5) (Garrison & Marth, 2012). Samples with median UMI count < 10 across MIPs were repooled and resequenced to obtain higher sequencing read depth. The VCF file generated after variant calling was used for downstream population genomic analyses. We leveraged the MIPLicorn R package (https://github.com/bailey-lab/miplicorn) to extract PfCRT and HBB genotypes for prevalence analysis using R studio (version4.4.3).

### Estimation of complexity of infection

We used targeted IBC2FULL SNPs for discrete COI calculation with *THE REAL McCOIL* package (version2.0) (Chang et al., 2017). The total number of Markov chain Monte Carlo was set to 2000 with 500 burn-in iterations. We included only SNPs with MAF ≥1% in this analysis after variant filtering out sites and samples with ≥ 10% and 20% genotype missingness, respectively. We calculated the prevalence of polygenomic infections (i.e samples with COI > 1) by region and plotted results on the map using *sf*, *ggspatial* and *rnaturalearth* packages in R. Continuous COI was computed with the *coiaf* R package using the same SNP set for our samples and historical samples from the Pf7 dataset (Paschalidis et al., 2022).

### Identity-by-descent analysis

The relatedness between samples was estimated by measuring pairwise IBD using the filtered SNP data obtained with the IBC2FULL MIP panel. We used the hmmIBD package (Schaffner et al., 2018) with a maximum number of fit iterations of 5 and >= 200 informative sites between sample pairs and assumed 0.1% genotyping error rate to detect IBD. Mean pairwise IBD scores within districts, between districts and were calculated and visualized using *ggplot2*, *sf*, *ggrepel, ggspatial* and *rnaturalearth* packages in R. IBD network analysis was performed using the *igraph* package. The *geosphere* package was used to estimate distances between sites based on their Global Positioning System (GPS) coordinates for correlation analysis with IBD.

#### Population structure analysis

For SNPs-based population structure analysis, PLINK was employed to calculate the pairwise variance-standardized genetic relationship matrix between samples on which we performed principal component analysis (PCA) using the PCA package in R. We used the *factoextra* package to visualize the results of the PCA. For microhaplotypes-based population structure in monogenomic samples (n=234), microhaplotype corresponding to genomic regions captured by IBC2FULL MIPs were extracted from the VCF file by concatenating genotype nucleotides. We obtained a distribution of microhaplotypes by sample which was then transformed into a genotype table containing two values; 0 and 2 indicating reference and alternate haplotypes, respectively. This table was used to calculate pairwise F_st_-based genetic distances using the *hierfstat* package after removing haplotypes that were missing in ≥ 90% of samples and samples lacking ≥80% of haplotypes. The genetic distance matrix was used for principal component calculations as described above for the SNPs-based method.

#### Analysis of selection signatures

We used IsoRelate’s iR statistics to analyze signatures of recent positive selection across the genome based on IBD. Shared IBD segments ≥ 50kb between all pairwise combinations of samples were detected based on at least 20 SNPs. We theoretically assumed 0.1% genotype error rate and the VCF was filtered to drop SNPs and samples with ≥10% and 20% genotype missingness, respectively, prior to the analysis. The IsoRelate function *getIBDiR* was used to estimate the pairwise iR statistics across the genome with and without sample stratification by site.

### Haplotype pattern analysis

We leveraged SNPs (MAF≥1%) targeted by the IBC2FULL panel within 45Kb and ∼300Kb regions flanking *pfcrt* and *acs8* to analyze haplotype patterns around these genes in monogenomic samples (n=130) randomly selected across sites. Although only monogenomic samples were included, we selected only major alleles to avoid ambiguity in the genotype calls at low sequencing depth loci. The SNP set extracted from the VCF was converted into a genotype table containing two values (0 = reference and 2= alternate) with samples in the rows and SNP positions in the columns. This genotype table was visualized using the *ComplexHeatmap* R package (https://jokergoo.github.io/ComplexHeatmap-reference/book/a-single-heatmap.html).

The complete-linkage clustering method was used to cluster haplotypes. Sample Pfsa1 and PfCRT genotype information was added as bar plots to the heatmap.

## Authors contribution

KN, JJJ, JAB and AAD designed and led the study. KN analyzed data and interpreted results. JJJ, JAB and AAD supervised the analysis and interpretation of the data. JJJ and JAB obtained primary funding for the molecular work. AAD obtained primary funding for the surveillance activities country-wide in Mali. AD, SS, AK, HD,MT, BD, MD and BK performed sample and clinical data collection and DNA extraction. AAD and MSS led and supervised the surveillance activities. VS, MC, AK supported the surveillance activities. AC and KN sequenced samples. KN wrote the primary draft of the manuscript. All authors contributed to the writing of the manuscript.

## Data Availability

All data produced in the present study are available upon reasonable request to the authors

## Acknowledgements

This work was supported by the Gates Foundation, United States of America (INV-023065, INV-037659 and INV-043733 to AAD). The opinions and views in this article are solely the authors’ and do not reflect any impressions of the funding body. This study was additionally supported by the National Institutes for Allergy and Infectious Diseases (R01AI156267 to JAB and JJJ; K24AI134990 to JJJ). The authors are grateful to Malaria Research and Reference Reagent Resource Center (MR4) for the laboratory strain DNA controls. The authors thank study participants and caregivers at all the 13 health districts.

## References

1. Allison, A. C. (1954). Protection afforded by sickle-cell trait against subtertian malareal infection. British Medical Journal, 1(4857), 290–294.

2. Amambua-Ngwa, A., Jeffries, D., Amato, R., Worwui, A., Karim, M., Ceesay, S., Nyang, H., Nwakanma, D., Okebe, J., Kwiatkowski, D., Conway, D. J., & D’Alessandro, U. (2018). Consistent signatures of selection from genomic analysis of pairs of temporal and spatial Plasmodium falciparum populations from The Gambia. Scientific Reports, 8(1), 9687.

3. Aydemir, O., Janko, M., Hathaway, N. J., Verity, R., Mwandagalirwa, M. K., Tshefu, A. K., Tessema, S. K., Marsh, P. W., Tran, A., Reimonn, T., Ghani, A. C., Ghansah, A., Juliano, J. J., Greenhouse, B. R., Emch, M., Meshnick, S. R., & Bailey, J. A. (2018). Drug-resistance and population structure of Plasmodium falciparum across the Democratic Republic of Congo using high-throughput molecular inversion probes. The Journal of Infectious Diseases, 218(6), 946–955.

4. Band, G., Leffler, E. M., Jallow, M., Sisay-Joof, F., Ndila, C. M., Macharia, A. W., Hubbart, C., Jeffreys, A. E., Rowlands, K., Nguyen, T., Gonçalves, S., Ariani, C. V., Stalker, J., Pearson, R. D., Amato, R., Drury, E., Sirugo, G., d’Alessandro, U., Bojang, K. A., … Kwiatkowski, D. P. (2022). Malaria protection due to sickle haemoglobin depends on parasite genotype. Nature, 602(7895), 106–111.

5. Beck, H. E., Zimmermann, N. E., McVicar, T. R., Vergopolan, N., Berg, A., & Wood, E. F. (2018). Present and future Köppen-Geiger climate classification maps at 1-km resolution. Scientific Data, 5(1), 180214.

6. Björkman, A., & Bhattarai, A. (2005). Public health impact of drug resistant Plasmodium falciparum malaria. Acta Tropica, 94(3), 163–169.

7. Chang, H.-H., Worby, C. J., Yeka, A., Nankabirwa, J., Kamya, M. R., Staedke, S. G., Dorsey, G., Murphy, M., Neafsey, D. E., Jeffreys, A. E., Hubbart, C., Rockett, K. A., Amato, R., Kwiatkowski, D. P., Buckee, C. O., & Greenhouse, B. (2017). THE REAL McCOIL: A method for the concurrent estimation of the complexity of infection and SNP allele frequency for malaria parasites. PLoS Computational Biology, 13(1), e1005348.

8. Cissoko, M., Magassa, M., Sanogo, V., Ouologuem, A., Sangaré, L., Diarra, M., Bationo, C. S., Dolo, M., Bah, M. D., Doumbia, S., Coulibaly, M. B., Traoré, D., Sidibé, B., Landier, J., Cissé, I., Sacko, M., Gaudart, J., & Sagara, I. (2022). Stratification at the health district level for targeting malaria control interventions in Mali. Scientific Reports, 12(1), 8271.

9. Cissoko, M., Sagara, I., Landier, J., Guindo, A., Sanogo, V., Coulibaly, O. Y., Dembélé, P., Dieng, S., Bationo, C. S., Diarra, I., Magassa, M. H., Berthé, I., Katilé, A., Traoré, D., Dessay, N., & Gaudart, J. (2022). Sub-national tailoring of seasonal malaria chemoprevention in Mali based on malaria surveillance and rainfall data. Parasites & Vectors, 15(1), 278.

10. Conrad, M. D., Asua, V., Garg, S., Giesbrecht, D., Niaré, K., Smith, S., Namuganga, J. F., Katairo, T., Legac, J., Crudale, R. M., Tumwebaze, P. K., Nsobya, S. L., Cooper, R. A., Kamya, M. R., Dorsey, G., Bailey, J. A., & Rosenthal, P. J. (2023). Evolution of Partial Resistance to Artemisinins in Malaria Parasites in Uganda. The New England Journal of Medicine, 389(8), 722–732.

11. da Silva, C., Boene, S., Datta, D., Rovira-Vallbona, E., Aranda-Díaz, A., Cisteró, P., Hathaway, N., Tessema, S., Chidimatembue, A., Matambisso, G., Nhama, A., Macete, E., Pujol, A., Nhamussua, L., Galatas, B., Guinovart, C., Enosse, S., De Carvalho, E., Rogier, E., … Mayor, A. (2023). Targeted and whole-genome sequencing reveal a north-south divide in P. falciparum drug resistance markers and genetic structure in Mozambique. Communications Biology, 6(1), 619.

12. Deutsch-Feldman, M., Aydemir, O., Carrel, M., Brazeau, N. F., Bhatt, S., Bailey, J. A., Kashamuka, M., Tshefu, A. K., Taylor, S. M., Juliano, J. J., Meshnick, S. R., & Verity, R. (2019). The changing landscape of Plasmodium falciparum drug resistance in the Democratic Republic of Congo. BMC Infectious Diseases, 19(1), 872.

13. de Vries, L. E., Jansen, P. A. M., Barcelo, C., Munro, J., Verhoef, J. M. J., Pasaje, C. F. A., Rubiano, K., Striepen, J., Abla, N., Berning, L., Bolscher, J. M., Demarta-Gatsi, C., Henderson, R. W. M., Huijs, T., Koolen, K. M. J., Tumwebaze, P. K., Yeo, T., Aguiar, A. C. C., Angulo-Barturen, I., … Dechering, K. J. (2022). Preclinical characterization and target validation of the antimalarial pantothenamide MMV693183. Nature Communications, 13(1), 2158.

14. Diarra, Y., Koné, O., Sangaré, L., Doumbia, L., Haidara, D. B. B., Diallo, M., Maiga, A., Sango, H. A., Sidibé, H., Mihigo, J., Nace, D., Ljolje, D., Talundzic, E., Udhayakumar, V., Eckert, E., Woodfill, C. J., Moriarty, L. F., Lim, P., Krogstad, D. J., … Koita, O. A. (2021). Therapeutic efficacy of artemether-lumefantrine and artesunate-amodiaquine for the treatment of uncomplicated Plasmodium falciparum malaria in Mali, 2015-2016. Malaria Journal, 20(1), 235.

15. Djimde, A. A., Barger, B., Kone, A., Beavogui, A. H., Tekete, M., Fofana, B., Dara, A., Maiga, H., Dembele, D., Toure, S., Dama, S., Ouologuem, D., Sangare, C. P. O., Dolo, A., Sogoba, N., Nimaga, K., Kone, Y., & Doumbo, O. K. (2010). A molecular map of chloroquine resistance in Mali. FEMS Immunology and Medical Microbiology, 58(1), 113–118.

16. Djimdé, A., Doumbo, O. K., Cortese, J. F., Kayentao, K., Doumbo, S., Diourté, Y., Coulibaly, D., Dicko, A., Su, X. Z., Nomura, T., Fidock, D. A., Wellems, T. E., & Plowe, C. V. (2001). A molecular marker for chloroquine-resistant falciparum malaria. The New England Journal of Medicine, 344(4), 257–263.

17. Duffy, C. W., Assefa, S. A., Abugri, J., Amoako, N., Owusu-Agyei, S., Anyorigiya, T., MacInnis, B., Kwiatkowski, D. P., Conway, D. J., & Awandare, G. A. (2015). Comparison of genomic signatures of selection on Plasmodium falciparum between different regions of a country with high malaria endemicity. BMC Genomics, 16(1), 527.

18. Garrison, E., & Marth, G. (2012). Haplotype-based variant detection from short-read sequencing. In arXiv [q-bio.GN]. arXiv. http://arxiv.org/abs/1207.3907

19. Hamilton, W. L., Band, G., Aniweh, Y., Asoala, V., Drury, E., Rockett, K. A., Awandare, G. A., Kwiatkowski, D. P., & Amenga-Etego, L. N. (2023). A fourth locus in the Plasmodium falciparum genome associated with sickle haemoglobin. In bioRxiv (p. 2023.09.14.557461). 10.1101/2023.09.14.557461

20. Hills, T., Srivastava, A., Ayi, K., Wernimont, A. K., Kain, K., Waters, A. P., Hui, R., & Pizarro, J. C. (2011). Characterization of a new phosphatase from Plasmodium. Molecular and Biochemical Parasitology, 179(2), 69–79.

21. Juliano, J. J., Giesbrecht, D. J., Simkin, A., Fola, A. A., Lyimo, B. M., Pereus, D., Bakari, C., Madebe, R. A., Seth, M. D., Mandara, C. I., Popkin-Hall, Z. R., Moshi, R., Mbwambo, R. B., Niaré, K., MacInnis, B., Francis, F., Mbwambo, D., Garimo, I., Chacky, F., … Ishengoma, D. S. (2023). Country wide surveillance reveals prevalent artemisinin partial resistance mutations with evidence for multiple origins and expansion of high level sulfadoxine-pyrimethamine resistance mutations in northwest Tanzania. medRxiv: The Preprint Server for Health Sciences. 10.1101/2023.11.07.23298207

22. Klein, E. Y. (2013). Antimalarial drug resistance: a review of the biology and strategies to delay emergence and spread. International Journal of Antimicrobial Agents, 41(4), 311–317.

23. Konaté, D., Diawara, S. I., Touré, M., Diakité, S. A. S., Guindo, A., Traoré, K., Diarra, A., Keita, B., Thiam, S., Keita, M., Sissoko, I., Sogoba, N., Traoré, S. F., Krogtad, D. J., Doumbia, S., & Diakité, M. (2020). Effect of routine seasonal malaria chemoprevention on malaria trends in children under 5 years in Dangassa, Mali. Malaria Journal, 19(1), 137.

24. Laufer, M. K., Thesing, P. C., Eddington, N. D., Masonga, R., Dzinjalamala, F. K., Takala, S. L., Taylor, T. E., & Plowe, C. V. (2006). Return of chloroquine antimalarial efficacy in Malawi. The New England Journal of Medicine, 355(19), 1959–1966.

25. Liu, D. Q., Liu, R. J., Ren, D. X., Gao, D. Q., Zhang, C. Y., Qui, C. P., Cai, X. Z., Ling, C. F., Song, A. H., & Tang, X. (1995). Changes in the resistance of Plasmodium falciparum to chloroquine in Hainan, China. Bulletin of the World Health Organization, 73(4), 483–486.

26. Lubell, Y., Dondorp, A., Guérin, P. J., Drake, T., Meek, S., Ashley, E., Day, N. P. J., White, N. J., & White, L. J. (2014). Artemisinin resistance – modelling the potential human and economic costs. Malaria Journal, 13(1), 452.

27. Mahamar, A., Sumner, K. M., Levitt, B., Freedman, B., Traore, A., Barry, A., Issiaka, D., Dembele, A. B., Kanoute, M. B., Attaher, O., Diarra, B. N., Sagara, I., Djimde, A., Duffy, P. E., Fried, M., Taylor, S. M., & Dicko, A. (2022). Effect of three years’ seasonal malaria chemoprevention on molecular markers of resistance of Plasmodium falciparum to sulfadoxine-pyrimethamine and amodiaquine in Ouelessebougou, Mali. Malaria Journal, 21(1), 39.

28. Maiga, H., Grivoyannis, A., Sagara, I., Traore, K., Traore, O. B., Tolo, Y., Traore, A., Bamadio, A., Traore, Z. I., Sanogo, K., Doumbo, O. K., Plowe, C. V., & Djimde, A. A. (2021). Selection of pfcrt K76 and pfmdr1 N86 coding alleles after uncomplicated malaria treatment by artemether-lumefantrine in Mali. International Journal of Molecular Sciences, 22(11), 6057.

29. Malaria GEN, Abdel Hamid, M. M., Abdelraheem, M. H., Acheampong, D. O., Ahouidi, A., Ali, M., Almagro-Garcia, J., Amambua-Ngwa, A., Amaratunga, C., Amenga-Etego, L., Andagalu, B., Anderson, T., Andrianaranjaka, V., Aniebo, I., Aninagyei, E., Ansah, F., Ansah, P. O., Apinjoh, T., Arnaldo, P., … van der Pluijm, R. W. (2023). Pf7: an open dataset of Plasmodium falciparum genome variation in 20,000 worldwide samples. Wellcome Open Research, 8, 22.

30. Moser, K. A., Madebe, R. A., Aydemir, O., Chiduo, M. G., Mandara, C. I., Rumisha, S. F., Chaky, F., Denton, M., Marsh, P. W., Verity, R., Watson, O. J., Ngasala, B., Mkude, S., Molteni, F., Njau, R., Warsame, M., Mandike, R., Kabanywanyi, A. M., Mahende, M. K., … Bailey, J. A. (2020). Describing the current status of Plasmodium falciparum population structure and drug resistance within mainland Tanzania using molecular inversion probes. Molecular Ecology. 10.1111/mec.15706

31. Niaré, K., Crudale, R., Fola, A. A., Wernsman Young, N., Asua, V., Conrad, M. D., Gashema, P., Ghansah, A., Hangi, S., Ishengoma, D. S., Mazarati, J.-B., Zeleke, A. J., Rosenthal, P. J., Djimdé, A. A., Juliano, J. J., & Bailey, J. A. (2025). Highly multiplexed molecular inversion probe panel in Plasmodium falciparum targeting common SNPs approximates whole-genome sequencing assessments for selection and relatedness. Frontiers in Genetics, 16, 1526049.

32. Paschalidis, A., Watson, O. J., Aydemir, O., Verity, R., & Bailey, J. A. (2022). coiaf: directly estimating complexity of infection with allele frequencies. In bioRxiv (p. 2022.05.26.493561). 10.1101/2022.05.26.493561

33. Python, A., Forster, A. J., Todd, P., Niare, K., Conrad, M., Ishengoma, D. S., Amenga-Etego, L. N., Lian, J., Liu, B., Yan, Y., Liang, J., Khan, M. M., Zheng, J., Bailey, J., Conway, D. J., Mentzer, A. J., Amambua-Ngwa, A., Leffler, E. M., Williams, T. N., & Band, G. (2025). Geographical variation drives adaptive equilibrium of the P. falciparum sickle-associated mutations. In bioRxiv (p. 2025.08.31.672853). 10.1101/2025.08.31.672853

34. Ranque, B., Kitenge, R., Ndiaye, D. D., Ba, M. D., Adjoumani, L., Traore, H., Coulibaly, C., Guindo, A., Boidy, K., Mbuyi, D., Ly, I. D., Offredo, L., Diallo, D. A., Tolo, A., Kafando, E., Tshilolo, L., & Diagne, I. (2022). Estimating the risk of child mortality attributable to sickle cell anaemia in sub-Saharan Africa: a retrospective, multicentre, case-control study. The Lancet. Haematology, 9(3), e208–e216.

35. Sangho, H., Keïta, H. D., Keïta, A. S., Diarra, F. Y., Belemou, B., Dia, A., Traoré, M., Keïta, F. D., Diarra, A., Diakité, B., Diallo, D., & Sidibé, T. (2009). Management of sickle cells disease by households in Bamako. Le Mali medical, 24(3), 53–56.

36. Schaffner, S. F., Badiane, A., Khorgade, A., Ndiop, M., Gomis, J., Wong, W., Ndiaye, Y. D., Diedhiou, Y., Thwing, J., Seck, M. C., Early, A., Sy, M., Deme, A., Diallo, M. A., Sy, N., Sene, A., Ndiaye, T., Sow, D., Dieye, B., … Volkman, S. K. (2023). Malaria surveillance reveals parasite relatedness, signatures of selection, and correlates of transmission across Senegal. Nature Communications, 14(1), 7268.

37. Schaffner, S. F., Taylor, A. R., Wong, W., Wirth, D. F., & Neafsey, D. E. (2018). hmmIBD: software to infer pairwise identity by descent between haploid genotypes. Malaria Journal, 17(1), 196.

38. Simmons, C. F., Gibbons, J., Zhang, M., Oberstaller, J., Pires, C. V., Casandra, D., Wang, C., Seyfang, A., Otto, T. D., Rayner, J. C., & Adams, J. H. (2023). Protein KIC5 is a novel regulator of artemisinin stress response in the malaria parasite Plasmodium falciparum. Scientific Reports, 13(1), 399.

39. Slater, H. C., Griffin, J. T., Ghani, A. C., & Okell, L. C. (2016). Assessing the potential impact of artemisinin and partner drug resistance in sub-Saharan Africa. Malaria Journal, 15(1), 10.

40. Taylor, S. M., Parobek, C. M., & Fairhurst, R. M. (2012). Haemoglobinopathies and the clinical epidemiology of malaria: a systematic review and meta-analysis. The Lancet Infectious Diseases, 12(6), 457–468.

41. Tessema, S. K., Hathaway, N. J., Teyssier, N. B., Murphy, M., Chen, A., Aydemir, O., Duarte, E. M., Simone, W., Colborn, J., Saute, F., Crawford, E., Aide, P., Bailey, J. A., & Greenhouse, B. (2022). Sensitive, Highly Multiplexed Sequencing of Microhaplotypes From the Plasmodium falciparum Heterozygome. The Journal of Infectious Diseases, 225(7), 1227–1237.

42. Tuteja, R. (2016). Emerging functions of helicases in regulation of stress survival in malaria parasite Plasmodium falciparum and their comparison with human host. Parasitology International, *65*(6 Pt A), 645–664.

43. Uwimana, A., Legrand, E., Stokes, B. H., Ndikumana, J.-L. M., Warsame, M., Umulisa, N., Ngamije, D., Munyaneza, T., Mazarati, J.-B., Munguti, K., Campagne, P., Criscuolo, A., Ariey, F., Murindahabi, M., Ringwald, P., Fidock, D. A., Mbituyumuremyi, A., & Menard, D. (2020). Emergence and clonal expansion of in vitro artemisinin-resistant Plasmodium falciparum kelch13 R561H mutant parasites in Rwanda. Nature Medicine, 26(10), 1602–1608.

44. Vanheer, L. N., Mahamar, A., Manko, E., Niambele, S. M., Sanogo, K., Youssouf, A., Dembele, A., Diallo, M., Maguiraga, S. O., Phelan, J., Osborne, A., Spadar, A., Smit, M. J., Bousema, T., Drakeley, C., Clark, T. G., Stone, W., Dicko, A., & Campino, S. (2023). Genome-wide genetic variation and molecular surveillance of drug resistance in Plasmodium falciparum isolates from asymptomatic individuals in Ouélessébougou, Mali. Scientific Reports, 13(1), 9522.

45. Veiga, M. I., Dhingra, S. K., Henrich, P. P., Straimer, J., Gnädig, N., Uhlemann, A.-C., Martin, R. E., Lehane, A. M., & Fidock, D. A. (2016). Globally prevalent PfMDR1 mutations modulate Plasmodium falciparum susceptibility to artemisinin-based combination therapies. Nature Communications, 7(1), 11553.

46. Venkatesan, M., Gadalla, N. B., Stepniewska, K., Dahal, P., Nsanzabana, C., Moriera, C., Price, R. N., Mårtensson, A., Rosenthal, P. J., Dorsey, G., Sutherland, C. J., Guérin, P., Davis, T. M. E., Ménard, D., Adam, I., Ademowo, G., Arze, C., Baliraine, F. N., Berens-Riha, N., … Asaq Molecular Marker Study Group. (2014). Polymorphisms in Plasmodium falciparum chloroquine resistance transporter and multidrug resistance 1 genes: parasite risk factors that affect treatment outcomes for P. falciparum malaria after artemether-lumefantrine and artesunate-amodiaquine. The American Journal of Tropical Medicine and Hygiene, 91(4), 833–843.

47. Verity, R., Aydemir, O., Brazeau, N. F., Watson, O. J., Hathaway, N. J., Mwandagalirwa, M. K., Marsh, P. W., Thwai, K., Fulton, T., Denton, M., Morgan, A. P., Parr, J. B., Tumwebaze, P. K., Conrad, M., Rosenthal, P. J., Ishengoma, D. S., Ngondi, J., Gutman, J., Mulenga, M., … Juliano, J. J. (2020). The impact of antimalarial resistance on the genetic structure of Plasmodium falciparum in the DRC. Nature Communications, 11(1), 2107.

48. World Health Organization. (2012). WHO policy recommendation: seasonal malaria chemoprevention (SMC) for plasmodium falciparum malaria control in highly seasonal transmission areas of the Sahel sub-region in Africa (No. WHO/HTM/GMP/2012.02). World Health Organization. https://iris.who.int/handle/10665/337978

49. World malaria report 2024. (n.d.). Retrieved January 17, 2025, from https://www.who.int/teams/global-malaria-programme/reports/world-malaria-report-2024

50. Young, N. W., Gashema, P., Giesbrecht, D., Munyaneza, T., Maisha, F., Mwebembezi, F., Budodo, R., Leonetti, A., Crudale, R., Iradukunda, V., Bosco, N. J., Boyce, R. M., Mandara, C. I., Kanyankole, G. K., Mulogo, E., Ishengoma, D. S., Hangi, S., Karema, C., Mazarati, J.-B., … Bailey, J. A. (2024). High frequency of artemisinin partial resistance mutations in the great lake region revealed through rapid pooled deep sequencing. In bioRxiv. 10.1101/2024.04.29.24306442

