## Supplemental materials for "Genome-wide SNP genotyping of *Plasmodium falciparum* isolates across Mali reveals major impacts of *Pfsa1* and PfCRT K76T selections on parasite populations"

**Table of content**

**SUPPLEMENTARY TABLES	1**

Table S1: Locations of samples retained for population genetics analyses	1

**SUPPLEMENTARY FIGURES	2**

Figure S1: Positive selective signatures by district.	2

Figure S2: Microhaplotypes-based PCA in mono-genomic samples (n=242) colored by district.	3

Figure S3: Microhaplotypes-based PCA in mono-genomic samples (n=242) colored by PfCRT genotype.	4

Figure S4: Comparison of the prevalences of PfCRT K76T mutation in samples carrying Pfsa1 versus wild type in historical samples.	5

Figure S5: Loading values of SNPs for the first dimension of the principal component analysis.	6

### **SUPPLEMENTARY TABLES**

#### **Table S1:** L**ocations of samples retained for population genetics analyses**

| **District** | **Region** | **Admin1** | **Sample size** |
| --- | --- | --- | --- |
| Bafoulabé | South-West | Kayes | 65 |
| Bamako | South | Bamako | 25 |
| Dioïla | South | Koulikoro | 67 |
| Diré | North | Tombouctou | 19 |
| Djenné | Central | Mopti | 26 |
| Gao | North | Gao | 7 |
| Kidal | North | Kidal | 5 |
| Kita | South-West | Kayes | 53 |
| Kolondiéba | South | Sikasso | 59 |
| Nioro | South-West | Kayes | 36 |
| Ségou | Central | Segou | 34 |
| Taoudeni | North | Tombouctou | 4 |
| Yélimané | South-West | Kayes | 10 |

### SUPPLEMENTARY FIGURES

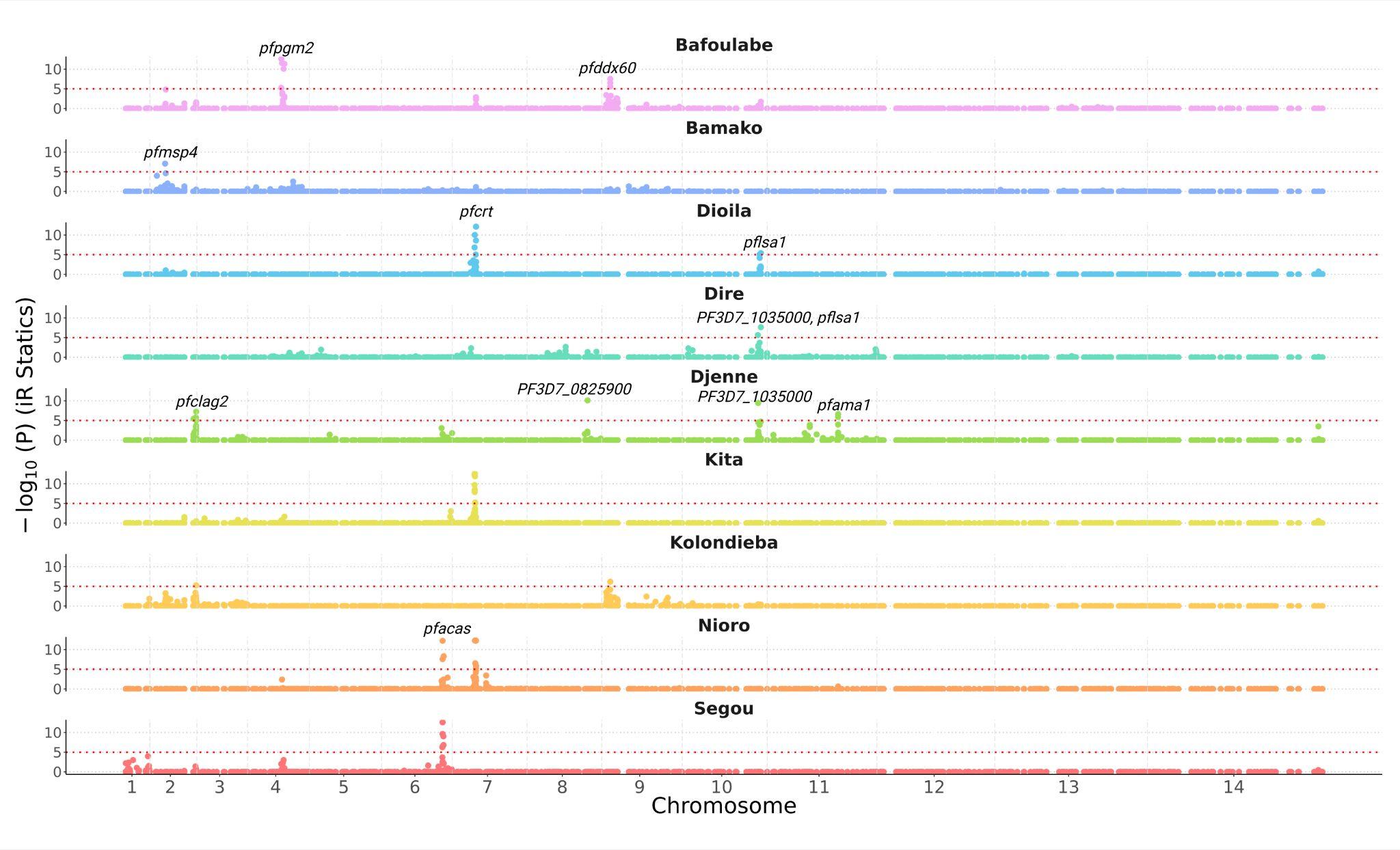

#### Figure S1: Positive selective signatures by district.

Data analysis included 410 samples collected in thirteen 10 health districts during the 2023 high malaria transmission. Samples from Dire (n=19) and Taoudeni (n=4) were merged and labeled as Dire. Y-axis represents the P value of the IsoRelate IR static obtained after estimating pairwise IBD between samples of the same district. P values were corrected for multiple testing using the false discovery rate. Signals with P value < 10^-5^ were considered significant. The names of genes found at the selective sweeps are indicated.

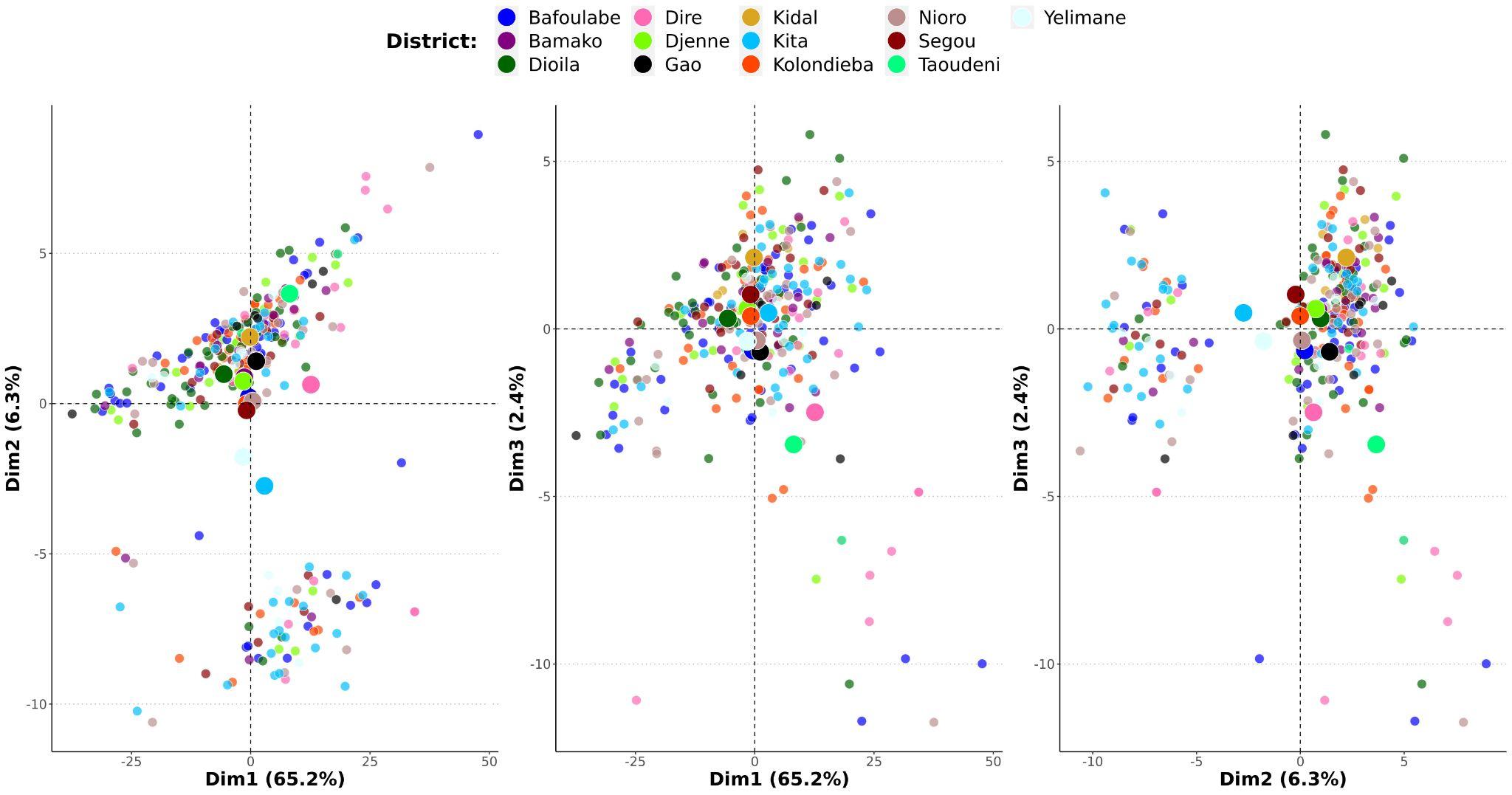

#### **Figure S2: Microhaplotypes-based PCA in mono-genomic samples (n=242) colored by district**.

First two dimensions (dim1 and 2) are shown and the percentage of variance explained by each of them is indicated in parentheses.

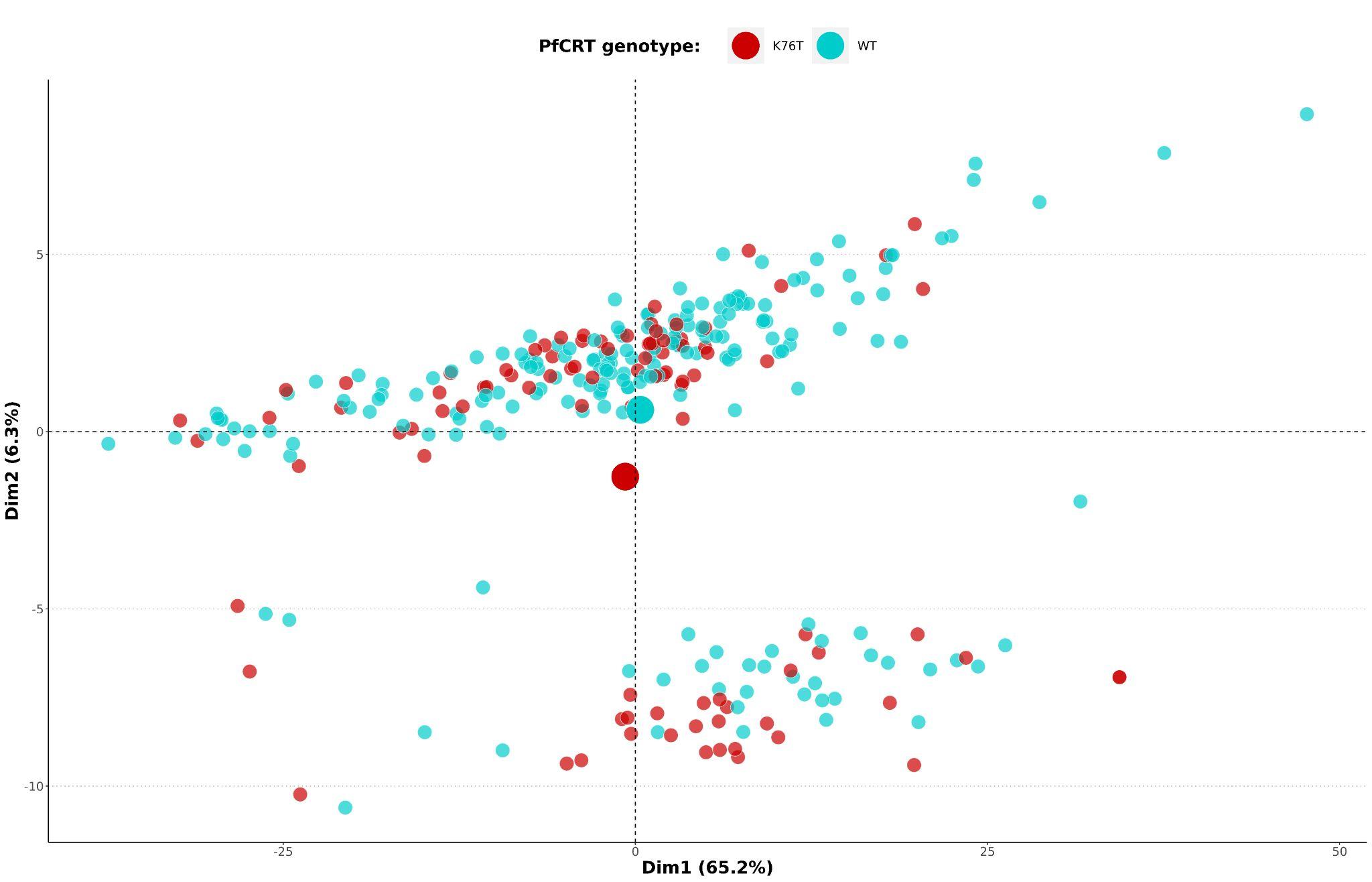

#### **Figure S3: Microhaplotypes-based PCA in mono-genomic samples (n=242) colored by PfCRT genotype**.

First two dimensions (dim1 and 2) are shown and the percentage of variance explained by each of them is indicated in parentheses. WT: wild type or K76.

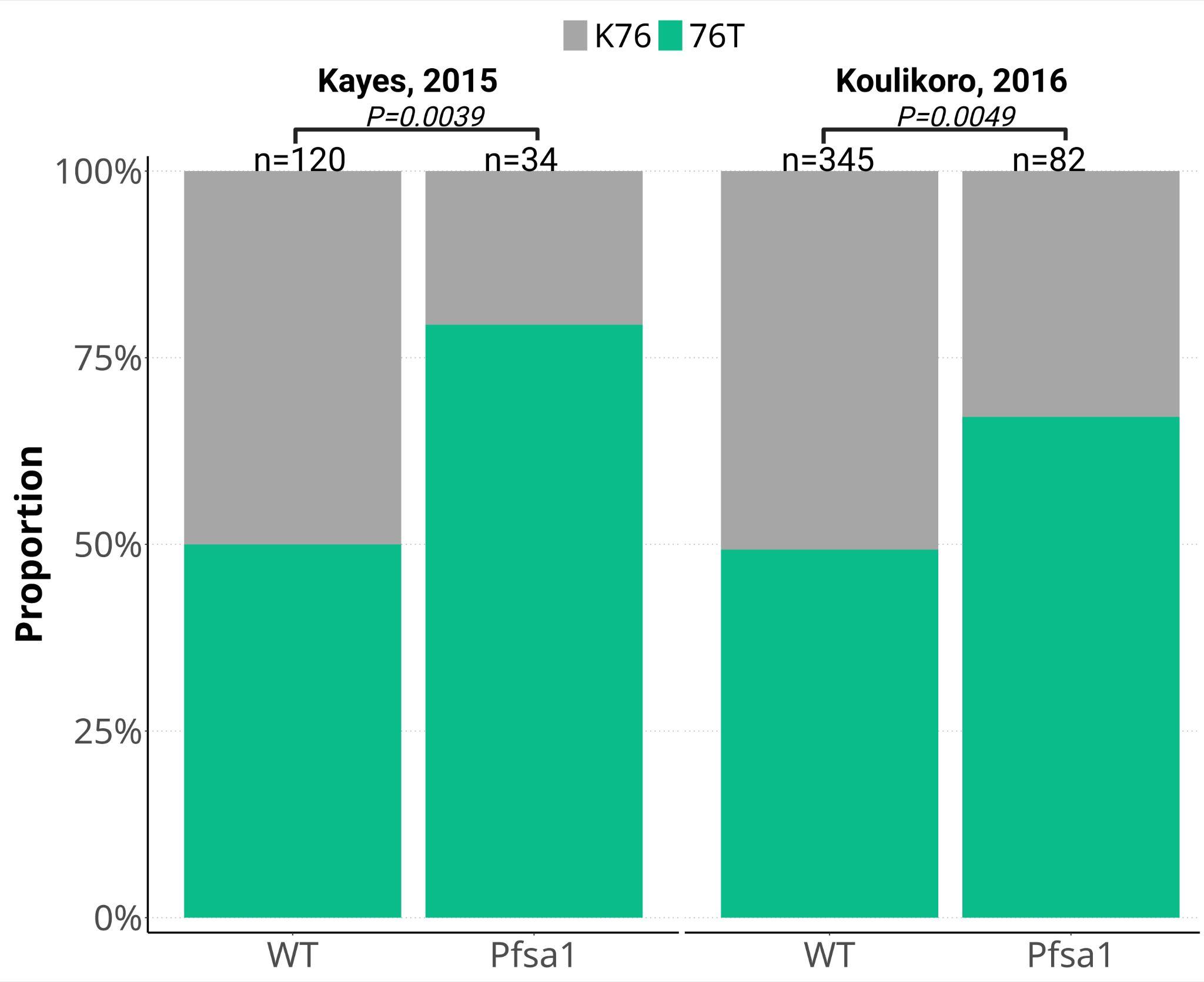

#### **Figure S4: Comparison of the prevalences of PfCRT K76T mutation in samples carrying *Pfsa1* versus wild type in historical samples**.

Samples from Kayes in 2015 and Koulikoro in 2016 were analyzed using the Pf7 dataset. Sample sizes and *P-*values (Chi-square test with Monte Carlo simulation using 100,000 replicates) are indicated.

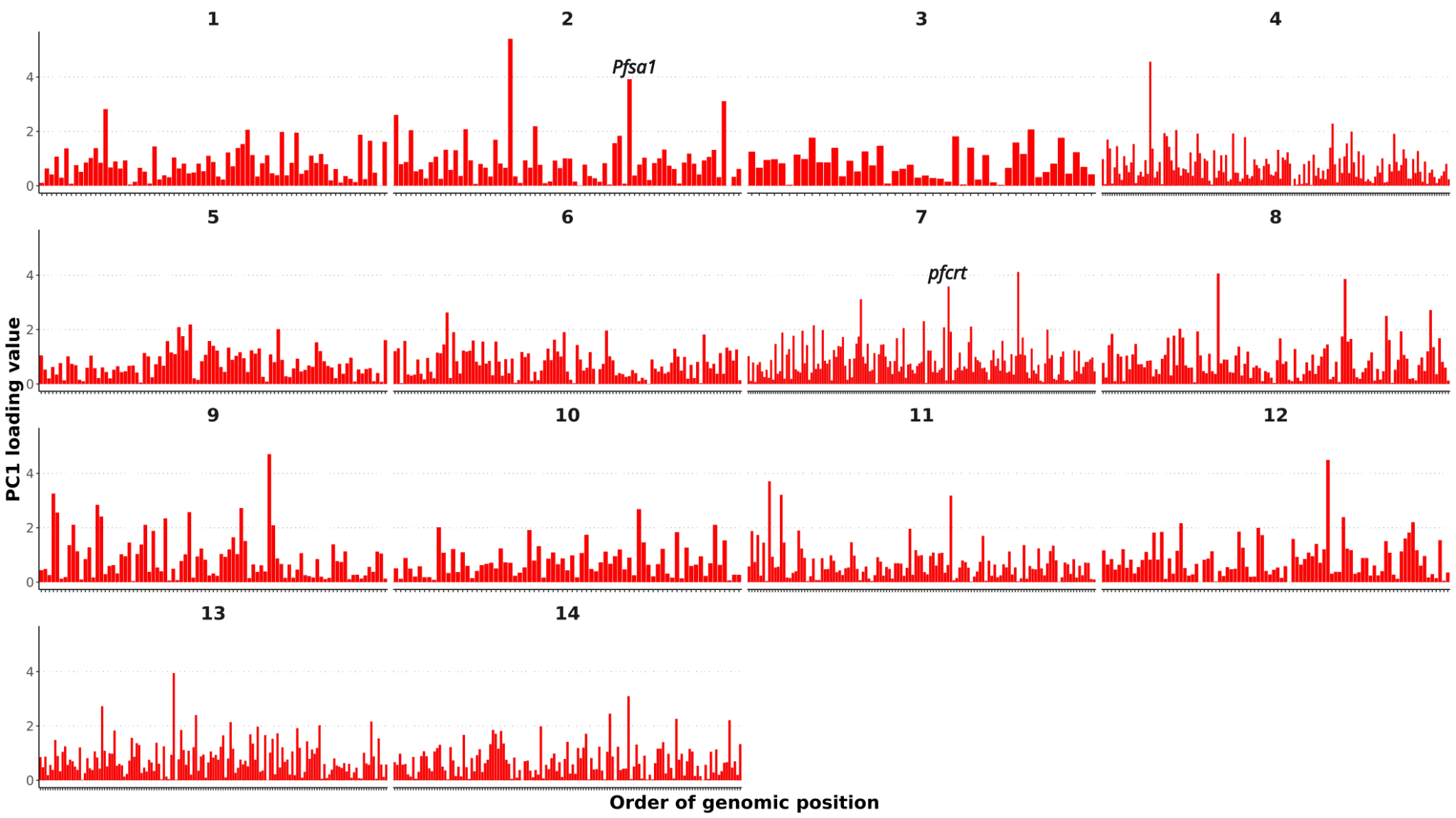

#### Figure S5: Loading values of SNPs for the first dimension of the principal component analysis.

Numbers on top indicate chromosomes. *Pfsa1* and *pfcrt* loci are marked. PC1: principal component 1.
